# Incidence of prostate cancer in Medicaid beneficiaries with and without HIV in 2001-2015 in 14 states

**DOI:** 10.1101/2024.05.24.24307676

**Authors:** Filip Pirsl, Keri Calkins, Jacqueline E. Rudolph, Eryka Wentz, Xiaoqiang Xu, Bryan Lau, Corinne E. Joshu

**Affiliations:** Department of Epidemiology, Johns Hopkins Bloomberg School of Public Health, Baltimore, Maryland, United States; Mathematica, Ann Arbor, Michigan, United States; Department of Psychiatry and Behavioral Sciences, Johns Hopkins School of Medicine, Baltimore, Maryland, United States; Department of Oncology, Sidney Kimmel Comprehensive Cancer Center at Johns Hopkins, Baltimore, Maryland, United States; Department of Medicine, Division of General Internal Medicine, Johns Hopkins School of Medicine, Baltimore, Maryland, United States

**Keywords:** HIV, prostate cancer, Medicaid, non-AIDS-defining cancer

## Abstract

**Background:** Prostate cancer is projected to be the most common cancer among people living with HIV; however, incidence of prostate cancer has been reported to be lower in men with HIV compared to men without HIV with little evidence to explain this difference. We describe prostate cancer incidence by HIV status in Medicaid beneficiaries, allowing for comparison of men with and without HIV who are similar with respect to socioeconomic characteristics and access to healthcare.

**Methods:** Medicaid beneficiaries (N=15,167,636) aged 18-64 with ≥7 months of continuous enrollment during 2001-2015 in 14 US states were retained for analysis. Diagnoses of HIV and prostate cancer were identified using inpatient and other non-drug claims. We estimated cause-specific (csHR) and sub-distribution hazard ratios comparing incidence of prostate cancer by HIV status, adjusted for age, race-ethnicity, state of residence, year of enrollment, and comorbid conditions. Models were additionally stratified by age and race-ethnicity.

**Results:** There were 366 cases of prostate cancer observed over 299,976 person-years among beneficiaries with HIV and 17,224 cases over 22,298,914 person-years in beneficiaries without HIV. The hazard of prostate cancer was lower in men with HIV than men without HIV (csHR=0.89; 95% CI: 0.80, 0.99), but varied by race-ethnicity, with similar observations among non-Hispanic Black (csHR=0.79; 95% CI: 0.69, 0.91) and Hispanic (csHR=0.85; 95% CI: 0.67, 1.09), but not non-Hispanic white men (csHR=1.17; 95% CI: 0.91, 1.50). Results were similar in models restricted to ages 50-64 and 40-49, except for a higher hazard of prostate cancer in Hispanic men with HIV in their 40s, while the hazard of prostate cancer was higher in men with HIV across all models for men aged 18-39.

**Conclusion:** Reported deficits in prostate cancer incidence by HIV status may be restricted to specific groups defined by age and race-ethnicity.

## Introduction

People living with HIV (PLWH) are projected to age considerably, with increasing morbidity and mortality due to age-related conditions, including cardiovascular disease, cognitive decline, and cancer, with the latter estimated to currently account for up to one third of deaths in this population (1–7). PLWH are at elevated risk for numerous cancers compared to the general United States (US) population, including cancers of the anus, cervix, liver, and lung as well as lymphoma and Kaposi sarcoma (1,8–11). Conversely, it has been reported that incidence of breast and prostate cancer is lower among PLWH than in the general population (8–14). Despite these observations, prostate cancer is projected to account for the greatest proportion of cancer burden among PLWH as this population continues to age (1).

Multiple differences exist between men with and without HIV that may contribute to the observed differences in incidence of prostate cancer. Several physiologic processes that have been reported to differ by HIV status, including testosterone metabolism and replacement (15–17), diabetes (18–21), and chronic inflammation (22), may influence pathophysiology of prostate cancer. Testosterone is believed to be necessary for development of prostate cancer, although neither testosterone deficiency nor replacement have been associated with prostate cancer (23–26). Diabetes has been associated with reduced risk of aggressive prostate cancer (27–29), and intraprostatic inflammation may promote development of prostate cancer (30), although it is unknown how HIV-related inflammation influences the prostate, if at all.

Differences by HIV status also exist in demographic characteristics that influence health outcomes. HIV disproportionately affects minority and socioeconomically disadvantaged communities (31,32), resulting in differences in racial composition and healthcare coverage by HIV status. In 2019, an estimated 40% of PLWH in the US were non-Hispanic Black individuals, who account for only 12% of the general population (33,34). Non-Hispanic Black men are also subject to numerous prostate cancer disparities, including greater incidence of any and advanced prostate cancer (35,36), lower likelihood of receiving appropriate treatment (37,38), and greater prostate cancer-specific mortality (39–41) compared to other men, although differences are attenuated in settings of equal access to care. Racial disparities in prostate cancer in the general population may influence differences observed by HIV status. Moreover, healthcare for approximately 40% of PLWH is covered by Medicaid compared to 13% of the general population, where private insurance is most common (70%) (42,43), reflecting socioeconomic differences which may affect access to or engagement with healthcare.

Lastly, prostate cancer screening is a significant driver of detection of prostate cancer and extent of screening influences observed incidence rates (44,45). Prostate cancer screening has been reported to be higher among privately-insured men with HIV (12) but not in lower income men with AIDS (13) compared to men without HIV. Current understanding is unable to explain lower incidence of prostate cancer in men with HIV, and the hypotheses described here may act in concert to yield these observations.

The objective of this study is to describe incidence of prostate cancer by HIV status in a large sample of Medicaid beneficiaries, allowing for comparison of men with HIV to men without HIV who are otherwise similar by socioeconomic characteristics and access to healthcare, effectively adjusting for these factors. We are also able to conduct comparisons stratified by race-ethnicity, which is implicated in multiple prostate cancer disparities in the general population, to describe any heterogeneity that exists by this characteristic. Findings of this study will complement the existing literature and contribute previously unreported descriptions of prostate cancer incidence accounting for key demographic characteristics of men with HIV, providing additional evidence for consideration of mechanisms underlying differences in prostate cancer incidence by HIV status.

## Methods

### Study Sample

Medicaid Analytic eXtract (MAX) data were obtained from the Centers for Medicare and Medicaid Services (CMS) for 2001-2015 for 14 US states. MAX data were available through 2015 for California, Georgia, New York, and Pennsylvania; through 2014 for Massachusetts, Ohio, Texas, and Washington; and through 2013 for Alabama, Colorado, Florida, Illinois, Maryland, and North Carolina. Data available for analysis included enrollment data and claims for inpatient care, outpatient care, and long-term therapy for beneficiaries aged 18-64. Beneficiaries with dual eligibility for Medicare or private insurance coverage were excluded to ensure all care received was captured in Medicaid claims. Enrollment was defined at the month level and beneficiaries were considered to be eligible in a month with at least 15 days of Medicaid coverage. Follow-up for states with MAX data through 2015 was administratively censored at the end of September 2015, immediately preceding the transition to ICD-10, and only ICD-9 codes were used to identify claims of interest.

Only beneficiaries’ first enrollment periods were considered, and beneficiaries could not re-enter and continue contributing person-time if re-enrollment was observed, as measures of interest could not be observed during a lapse in coverage. A six-month run-in period of continuous enrollment was implemented to exclude prevalent cases of any cancer, meaning beneficiaries needed at least seven months of continuous enrollment and to be cancer-free to be included in this analysis. The Johns Hopkins Bloomberg School of Public Health Institutional Review Board determined that this study represented a secondary analysis of existing Medicaid claims data and that it met criteria for exempt status.

### Study Measures

HIV and covariates of interest were measured during the run-in period and tabulated at the start of the seventh month of continuous enrollment, henceforth referred to as baseline. Age, race-ethnicity, sex, and state of residence were obtained from the MAX personal summary file. Race-ethnicity was categorized as non-Hispanic white, non-Hispanic Black, Hispanic, and other or unknown race-ethnicity. Only beneficiaries for whom male sex was recorded were retained for this analysis. State was defined at baseline and fixed.

Men with HIV were identified using a modified version of the CMS chronic condition definition for HIV of at least one inpatient claim or at least two long-term or other care claims within a one-year period with ICD-9 code indicating diagnosis of HIV (042, 042.X, 079.53, 795.71, V08). The service start date of the first claim was used as the date of diagnosis of HIV. Prostate cancer, the outcome of interest, was identified using a similar algorithm, with ICD-9 codes indicating prostate cancer diagnosis (185.X, V10.46). We also identified diagnoses of comorbid conditions, specifically those included in the Charlson comorbidity index (48–50). Codes used to identify the conditions are provided in Supplemental Materials.

### Statistical Analysis

Men contributed follow-up time at risk beginning on the first day of the seventh month of continuous enrollment in Medicaid until earliest of diagnosis of prostate cancer, diagnosis of any other cancer, age 65, lapse in enrollment, recorded date of death, or end of data coverage for state of residence. We calculated incidence rates of prostate cancer per 100,000 person-years at risk, stratified by age and race-ethnicity. Age-stratified analyses used cutoffs of ages 40 and 50 years. The United States Preventive Services Task Force recommended initiation of prostate cancer screening at age 50 for most of the study period (51–53), whereas the American Cancer Society had suggested initiation of screening in the 40s for high-risk men, namely Black men and men with family history of prostate cancer (54–56). For age-stratified analyses, person-time contributed was additionally constrained by aging into and out of age strata.

Cox proportional hazard models were used to estimate the cause-specific hazard ratio (csHR) of prostate cancer in men with HIV compared to men without HIV. Due to the considerable difference in risk of death due to any cause by HIV status, we also estimated sub-distribution hazard ratios (sdHR) using a Fine and Gray model, to compare incidence of prostate cancer by HIV status allowing for death as a competing risk to occur. Lastly, we estimated and plotted cumulative incidence curves of prostate cancer by HIV status and 95% confidence intervals using the Aalen-Johansen estimator and Greenwood’s formula, respectively. All models used age as the time scale. The distribution of age in men with HIV differs from that of men without HIV in the general population (1,46) as well as men without HIV enrolled in Medicaid (47), requiring careful adjustment for age in analyses of cancer incidence in this population. Using age as the time scale is one way to control for these differences. HRs were estimated in all men, omitting men of other or unknown race-ethnicity, and separately among non-Hispanic white, non-Hispanic Black, and Hispanic men.; We additionally ran models stratified by age (18-39, 40-49, and 50-64). All models were adjusted for race-ethnicity, state of residence, calendar year of enrollment, and comorbid conditions (none, one, or two or more). All hazard ratio estimates presented henceforth are from adjusted models.

## Results

Men with HIV were older at baseline (median age 43 (IQR: 36, 50) vs. 30 (IQR: 20, 44)), were more frequently non-Hispanic Black (43.1% vs. 18.2%), and had more often been diagnosed with at least one comorbid condition (32.0% vs. 10.7%) than men without HIV (**Table 1**). Over one quarter of men with HIV (26.2%) were aged 50 or older at enrollment compared to 16.7% of men without HIV.

**Table 1:** Beneficiary Characteristics at Baseline.

| Characteristic | HIV<br>N=112,498 | No HIV<br>N=15,055,138 |
| --- | --- | --- |
| Enrollment year |  |  |
| 2001 or earlier | 30.4% | 13.2% |
| 2002 | 5.9% | 6.0% |
| 2003 | 5.0% | 5.5% |
| 2004 | 4.6% | 5.5% |
| 2005 | 4.3% | 5.2% |
| 2006 | 3.8% | 4.8% |
| 2007 | 3.3% | 4.7% |
| 2008 | 4.4% | 5.6% |
| 2009 | 5.0% | 6.6% |
| 2010 | 5.0% | 6.8% |
| 2011 | 4.8% | 6.9% |
| 2012 | 4.4% | 6.8% |
| 2013 | 4.0% | 6.3% |
| 2014 | 13.0% | 13.1% |
| 2015 | 2.0% | 3.0% |
| Age |  |  |
| Median (IQR) | 43 (36, 50) | 30 (20, 44) |
| 18-24 | 5.7% | 37.1% |
| 25-29 | 6.3% | 11.7% |
| 30-34 | 10.3% | 10.2% |
| 35-39 | 15.8% | 9.0% |
| 40-44 | 18.6% | 8.1% |
| 45-49 | 17.1% | 7.1% |
| 50-54 | 12.1% | 6.3% |
| 55-59 | 6.8% | 5.2% |
| 60-64 | 7.3% | 5.3% |
| Race-ethnicity |  |  |
| Non-Hispanic white | 27.0% | 33.7% |
| Non-Hispanic Black | 43.1% | 18.2% |
| Hispanic | 20.5% | 33.1% |
| Other/Unknown | 9.4% | 15.0% |
| State |  |  |
| Alabama | 0.8% | 1.0% |
| California | 21.7% | 43.4% |
| Colorado | 0.5% | 0.9% |
| Florida | 10.4% | 4.3% |
| Georgia | 3.9% | 2.2% |
| Illinois | 4.5% | 4.3% |
| Maryland | 3.5% | 2.1% |
| Massachusetts | 3.4% | 3.4% |
| New York | 35.1% | 19.0% |
| North Carolina | 3.5% | 2.4% |
| Ohio | 2.9% | 5.0% |
| Pennsylvania | 2.9% | 4.9% |
| Texas | 4.8% | 4.0% |
| Washington | 2.2% | 3.1% |
| Comorbid condition |  |  |
| 0 | 68.0% | 89.3% |
| 1 | 22.2% | 7.5% |
| 2+ | 9.8% | 3.2% |

Baseline defined as first day of seventh month of enrollment following six-month run-in period; characteristics tabulated during run-in period; comorbid conditions considered listed in Supplemental Materials; abbreviations: IQR-interquartile range

We observed 366 cases of prostate cancer over 299,976 person-years among men with HIV and 17,224 cases over 22,298,914 person-years in men without HIV. Prostate cancer cases were predominantly observed among men aged 50-64, followed by men aged 40-49 and 18-39, and more frequently observed among non-Hispanic Black men, followed by Hispanic and non-Hispanic white men.

The cause-specific hazard of prostate cancer was lower among men with HIV compared to men without HIV (csHR=0.89; 95% CI: 0.80, 0.99); however, this association varied by age and race-ethnicity (**Figure 1**, **Table 2**). The hazard of prostate cancer was lower among non-Hispanic Black (csHR=0.79; 95% CI: 0.69, 0.91) and to a lesser extent among Hispanic (csHR=0.85; 95% CI: 0.67, 1.09) men with HIV compared to men without HIV. Conversely, non-Hispanic white men with HIV had a slightly higher (but not significantly different) hazard of prostate cancer compared to non-Hispanic White men without HIV (csHR=1.17; 95% CI: 0.91, 1.50). Results were consistent when estimating the sdHR, with minimal changes in estimates (**Table 2**).

**Figure 1:**
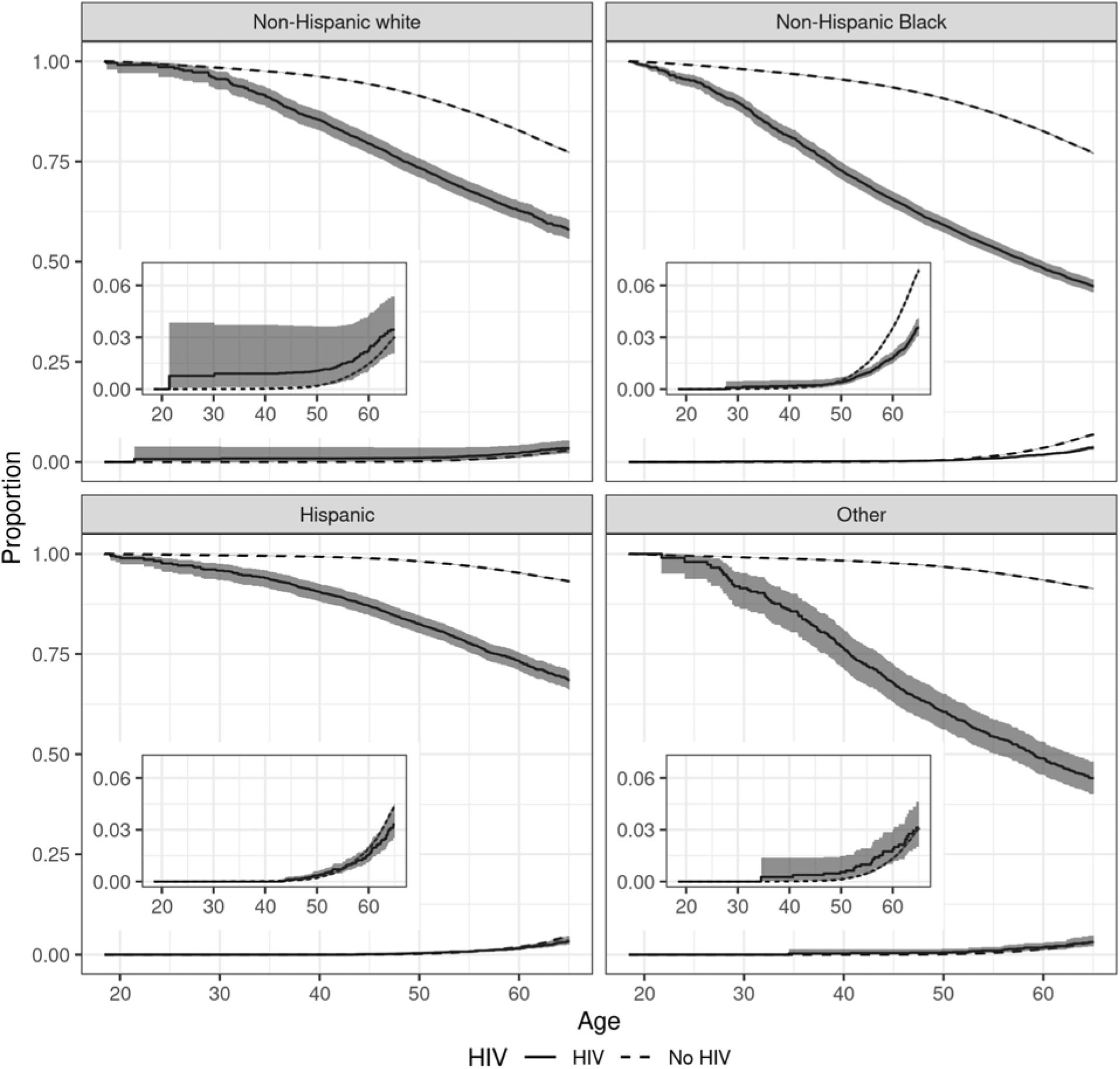
Cumulative Incidence of Prostate Cancer and Overall Survival by HIV Status in Men Aged 18-64 Stratified by Race-Ethnicity; insets are magnified curves for cumulative incidence of prostate cancer

**Table 2:** Cause-Specific, Sub-Distribution Hazard Ratios of Prostate Cancer by HIV Status in Men Aged 18-64.

|  | Cases | Person-time (PY) | IR<sup>a</sup> | Beneficiaries | Crude<sup>b</sup> |  | Adjusted<sup>c</sup> |  |
| --- | --- | --- | --- | --- | --- | --- | --- | --- |
|  |  |  |  |  | csHR (95% CI) | sdHR (95% CI) | csHR (95% CI) | sdHR (95% CI) |
| Overall |  |  |  |  |  |  |  |  |
| No HIV | 17,224 | 22,298,914.31 | 77.24 | 15,055,138 | 1.00 (reference) | 1.00 (reference) | 1.00 (reference) | 1.00 (reference) |
| HIV | 382 | 299,976.15 | 127.34 | 112,498 | 1.23 (1.11, 1.36) | 1.19 (1.08, 1.32) | 0.89 (0.80, 0.99) | 0.87 (0.87, 0.96) |
| NHW, NHB, Hispanic |  |  |  |  |  |  |  |  |
| No HIV | 15,130 | 19,014,049.38 | 79.57 | 12,798,610 | 1.00 (reference) | 1.00 (reference) | 1.00 (reference) | 1.00 (reference) |
| HIV | 357 | 281,072.75 | 127.01 | 101,941 | 1.14 (1.03, 1.26) | 1.11 (1.00, 1.23) | 0.85 (0.77, 0.95) | 0.83 (0.75, 0.92) |
| Non-Hispanic white |  |  |  |  |  |  |  |  |
| No HIV | 5,333 | 8,168,577.87 | 65.29 | 5,076,975 | 1.00 (reference) | 1.00 (reference) | 1.00 (reference) | 1.00 (reference) |
| HIV | 64 | 72,883.66 | 87.81 | 30,324 | 1.21 (0.95, 1.55) | 1.19 (0.93, 1.52) | 1.17 (0.91, 1.50) | 1.14 (0.89, 1.46) |
| Non-Hispanic Black |  |  |  |  |  |  |  |  |
| No HIV | 6,826 | 4,614,123.25 | 147.94 | 2,736,867 | 1.00 (reference) | 1.00 (reference) | 1.00 (reference) | 1.00 (reference) |
| HIV | 226 | 136,837.93 | 165.16 | 48,503 | 0.83 (0.73, 0.95) | 0.80 (0.70, 0.91) | 0.79 (0.69, 0.91) | 0.76 (0.67, 0.87) |
| Hispanic |  |  |  |  |  |  |  |  |
| No HIV | 2,971 | 6,231,348.25 | 47.68 | 4,984,768 | 1.00 (reference) | 1.00 (reference) | 1.00 (reference) | 1.00 (reference) |
| HIV | 67 | 71,351.16 | 93.90 | 23,114 | 1.05 (0.83, 1.34) | 1.03 (0.81, 1.32) | 0.85 (0.67, 1.09) | 0.83 (0.65, 1.07) |

<sup>a</sup>per 100,000 person-years; <sup>b</sup>adjusted for age (time metric), <sup>c</sup>adjusted for age (time metric), race-ethnicity, state of residence, enrollment year, and comorbid conditions; abbreviations: CI-confidence interval, csHR-cause-specific hazard ratio, IR-incidence rate, NHB-non-Hispanic Black, NHW-non-Hispanic white, PY-person-years, sdHR-sub-distribution hazard ratio

In analyses stratified by age, similar results were observed among men aged 50-64 to those estimated among men aged 18-64, as most events occurred in this age category (**Table 3**, **Supplemental Figure 1**). Overall, the hazard of prostate cancer was lower among men with HIV (csHR=0.89; 95% CI: 0.80, 0.99). When stratified by race-ethnicity, the hazard of prostate cancer was lower among non-Hispanic Black (adjusted csHR=0.81; 95% CI: 0.70, 0.94) and Hispanic (csHR=0.80; 95% CI: 0.61, 1.06), but not non-Hispanic white (csHR=1.15; 95% CI: 0.88, 1.50), men with HIV compared to men without HIV (**Table 3**). The sdHR results remained similar.

**Table 3:** Cause-Specific, Sub-Distribution Hazard Ratios of Prostate Cancer by HIV Status in Men Aged 50-64.

|  | Cases | Person-time (PY) | IR<sup>a</sup> | Beneficiaries | Crude<sup>b</sup> |  | Adjusted<sup>c</sup> |  |
| --- | --- | --- | --- | --- | --- | --- | --- | --- |
|  |  |  |  |  | csHR (95% CI) | sdHR (95% CI) | acsHR (95% CI) | asdHR (95% CI) |
| Overall |  |  |  |  |  |  |  |  |
| No HIV | 16,007 | 5,418,742.81 | 295.40 | 2,945,505 | 1.00 (reference) | 1.00 (reference) | 1.00 (reference) | 1.00 (reference) |
| HIV | 332 | 108,867.34 | 304.96 | 41,603 | 1.22 (1.09, 1.36) | 1.17 (1.05, 1.31) | 0.89 (0.80, 0.99) | 0.85 (0.77, 0.95) |
| NHW, NHB, Hispanic |  |  |  |  |  |  |  |  |
| No HIV | 14,012 | 4,446,608.25 | 315.12 | 2,341,829 | 1.00 (reference) | 1.00 (reference) | 1.00 (reference) | 1.00 (reference) |
| HIV | 311 | 102,146.14 | 304.47 | 37,923 | 1.14 (1.01, 1.27) | 1.09 (0.98, 1.22) | 0.85 (0.76, 0.96) | 0.82 (0.73, 0.92) |
| Non-Hispanic white |  |  |  |  |  |  |  |  |
| No HIV | 4,974 | 2,233,446.25 | 222.71 | 1,153,935 | 1.00 (reference) | 1.00 (reference) | 1.00 (reference) | 1.00 (reference) |
| HIV | 55 | 25,097.74 | 219.14 | 10,732 | 1.19 (0.91, 1.55) | 1.16 (0.89, 1.52) | 1.15 (0.88, 1.50) | 1.12 (0.86, 1.46) |
| Non-Hispanic Black |  |  |  |  |  |  |  |  |
| No HIV | 6,308 | 1,228,129.00 | 513.63 | 559,478 | 1.00 (reference) | 1.00 (reference) | 1.00 (reference) | 1.00 (reference) |
| HIV | 202 | 52,880.52 | 381.99 | 18,890 | 0.85 (0.74, 0.98) | 0.81 (0.70, 0.93) | 0.81 (0.70, 0.94) | 0.77 (0.67, 0.89) |
| Hispanic |  |  |  |  |  |  |  |  |
| No HIV | 2,730 | 985,033.00 | 277.15 | 628,416 | 1.00 (reference) | 1.00 (reference) | 1.00 (reference) | 1.00 (reference) |
| HIV | 54 | 24,167.88 | 223.44 | 8,301 | 0.97 (0.74, 1.27) | 0.94 (0.72, 1.23) | 0.80 (0.61, 1.06) | 0.78 (0.59, 1.02) |

<sup>a</sup>per 100,000 person-years, <sup>b</sup>adjusted for age (time metric), <sup>c</sup>adjusted for age (time metric), race-ethnicity, state of residence, enrollment year, and comorbid conditions; abbreviations: CI-confidence interval, csHR-cause-specific hazard ratio, IR-incidence rate, NHB-non-Hispanic Black, NHW-non-Hispanic white, PY-person-years, sdHR-sub-distribution hazard ratio

Among men aged 40-49, hazard of prostate cancer was also lower among men with HIV compared to men without HIV; however, this difference was attenuated compared to men aged over 50 (csHR=0.92; 95% CI: 0.68, 1.26) (**Table 4**, **Supplemental Figure 2**). When stratified by race-ethnicity, men with HIV had a lower hazard of prostate cancer compared to men without HIV among non-Hispanic Black men (csHR=0.68; 95% CI: 0.43, 1.06), while the hazard of prostate cancer was higher among men with HIV compared to men without HIV among non-Hispanic white (csHR=1.12; 95% CI: 0.53, 2.37) and Hispanic men (csHR=1.24; 95% CI: 0.69, 2.21). None of the differences by HIV status in this age group were statistically significant. No changes in results were observed when estimating the sdHR.

**Table 4:** Cause-Specific, Sub-Distribution Hazard Ratios of Prostate Cancer by HIV Status in Men Aged 40-49.

|  | Cases | Person-time (PY) | IR<sup>a</sup> | Beneficiaries | Crude<sup>b</sup> |  | Adjusted<sup>c</sup> |  |
| --- | --- | --- | --- | --- | --- | --- | --- | --- |
|  |  |  |  |  | csHR (95% CI) | sdHR (95% CI) | csHR (95% CI) | sdHR (95% CI) |
| Overall |  |  |  |  |  |  |  |  |
| No HIV | 1,062 | 4,376,172.00 | 24.27 | 2,736,466 | 1.00 (reference) | 1.00 (reference) | 1.00 (reference) | 1.00 (reference) |
| HIV | 43 | 116,470.90 | 36.92 | 49,401 | 1.43 (1.05, 1.93) | 1.39 (1.03, 1.89) | 0.92 (0.68, 1.26) | 0.90 (0.66, 1.23) |
| NHW, NHB, Hispanic |  |  |  |  |  |  |  |  |
| No HIV | 973 | 3,656,230.88 | 26.61 | 2,276,711 | 1.00 (reference) | 1.00 (reference) | 1.00 (reference) | 1.00 (reference) |
| HIV | 40 | 109,660.72 | 36.48 | 45,392 | 1.28 (0.93, 1.75) | 1.25 (0.91, 1.71) | 0.88 (0.64, 1.21) | 0.86 (0.62, 1.19) |
| Non-Hispanic white |  |  |  |  |  |  |  |  |
| No HIV | 312 | 1,672,119.12 | 18.66 | 982,198 | 1.00 (reference) | 1.00 (reference) | 1.00 (reference) | 1.00 (reference) |
| HIV | 7 | 29,985.88 | 23.34 | 13,768 | 1.23 (0.58, 2.59) | 1.20 (0.57, 2.54) | 1.12 (0.53, 2.37) | 1.09 (0.52, 2.31) |
| Non-Hispanic Black |  |  |  |  |  |  |  |  |
| No HIV | 456 | 883,193.50 | 51.63 | 490,435 | 1.00 (reference) | 1.00 (reference) | 1.00 (reference) | 1.00 (reference) |
| HIV | 20 | 51,005.41 | 39.21 | 21,177 | 0.72 (0.46, 1.13) | 0.71 (0.45, 1.10) | 0.68 (0.43, 1.06) | 0.66 (0.42, 1.03) |
| Hispanic |  |  |  |  |  |  |  |  |
| No HIV | 205 | 1,100,918.25 | 18.62 | 804,078 | 1.00 (reference) | 1.00 (reference) | 1.00 (reference) | 1.00 (reference) |
| HIV | 13 | 28,669.43 | 45.34 | 10,447 | 2.15 (1.22, 3.76) | 2.12 (1.21, 3.71) | 1.24 (0.69, 2.21) | 1.22<sup>c</sup> |

<sup>a</sup>per 100,000 person-years, <sup>b</sup>adjusted for age (time metric), <sup>c</sup>adjusted for age (time metric), race-ethnicity, state of residence, enrollment year, and comorbid conditions, <sup>c</sup>unable to estimate standard error and confidence interval; abbreviations: CI-confidence interval, csHR-cause-specific hazard ratio, IR-incidence rate, NHB-non-Hispanic Black, NHW-non-Hispanic white, PY-person-years, sdHR-sub-distribution hazard ratio

Lastly, very few prostate cancer cases (n=7) were observed among men aged 18-39, resulting in imprecise model estimates. Overall, the hazard of prostate cancer was elevated among men with HIV compared to men without HIV (csHR=2.02; 95% CI: 0.93, 4.39), with similar results observed among non-Hispanic Black (csHR=1.81; 95% CI: 0.64, 5.10) and non-Hispanic white men (csHR= csHR=4.15; 95% CI: 0.99, 17.43) (**Supplemental Table 1, Supplemental Figure 3**). Due to the small number of cases, a Cox model could not be run among Hispanic men, nor could we run the Fine-Gray model among any race-ethnicity group.

## Discussion

Men with HIV were found to have lower hazard of prostate cancer; however, heterogeneity in this association was observed by age and race-ethnicity. Among men aged 40 or older, the hazard of prostate cancer was largely lower in non-Hispanic Black, but higher in non-Hispanic white men with HIV compared to men without HIV. The relative hazard of prostate cancer by HIV status in Hispanic men varied by age category. Our results are congruent with previous reports of lower incidence of prostate cancer in men with HIV compared to men without HIV (8–14), although the estimates we report indicate a smaller difference. We additionally report model estimates stratified by race-ethnicity and describe previously unreported differences in the association between HIV and incidence of prostate cancer by race-ethnicity.

Differences observed by HIV status and race-ethnicity may be explained in part by prostate cancer screening, a significant driver of the detection of prostate cancer (44, 45). For most of the study period, guidelines recommended screening for men aged 50 or older, with some guidelines suggesting initiation of screening in the 40s for Black men and men with family history of prostate cancer (51–56). It is possible that, among men in whom screening is not yet recommended, men with HIV are more likely to be screened and have prostate cancer detected due to increased engagement in HIV care. At ages for which screening is recommended, this difference due to care engagement is abrogated or even reversed by broad uptake of screening in men without HIV. The recommendation of earlier initiation of screening in Black men may contribute to a lower hazard of prostate cancer observed in non-Hispanic Black men with HIV beginning already in men in their 40s. In contrast, among Hispanic men with HIV, the hazard of prostate cancer was higher in the 40s, but lower beyond age 50, than in men without HIV. This explanation is not supported by findings in non-Hispanic white men, in whom the hazard of prostate cancer was consistently comparable by HIV status across age categories, although this may reflect different patterns of screening uptake or cancer detection in either or both HIV groups compared to non-Hispanic Black and Hispanic men.

Biological mechanisms may explain the reduced incidence of prostate cancer in men with HIV compared to men without HIV, such as those mediated by inflammatory processes or hormonal perturbations. However, further refinements may be needed to explain the heterogeneity in associations by race-ethnicity observed in this analysis. It is possible that prostate cancer disparities by race-ethnicity described in the general population exert an influence on and potentially interact with biological processes and characteristics that differ by HIV status, although what these interactions are is unclear.

This study was not without limitations, and findings of these analyses should be interpreted with the following considerations. This analysis was conducted in men aged 18-64 whereas approximately 60% of prostate cancer cases in the US are diagnosed in men aged 65 or older (57). However, the proportion of PLWH in the US aged 65 or older was below 10% for much of the study period and only in more recent years have greater numbers of PLWH aged beyond 65 years (1). Nevertheless, the findings of this study are not generalizable to individuals aged 65 and older in whom prostate cancer burden is highest.

The outcome of interest in this study was incidence of any prostate cancer, as neither stage nor grade at diagnosis could be determined from administrative claims data. Results of this analysis should be interpreted solely in the context of total prostate cancer. Prostate cancer-specific mortality in the first five years following diagnosis increases with grade and almost exclusively occurs among men with regional or distant disease at diagnosis. While prostate cancer is most commonly diagnosed at the local stage, (57,58) men with HIV have previously been found to be diagnosed with prostate cancer at more advanced stages (59,60), which are associated with adverse outcomes.

A six-month run-in period was implemented to exclude beneficiaries with prevalent cancer at enrollment, and only beneficiaries with evidence of HIV observed during this period were considered to have HIV for analysis. Inclusion of beneficiaries diagnosed with HIV later in enrollment in analysis as not having HIV likely attenuated our estimates, assuming these individuals experienced risks indicated by our results after being diagnosed with HIV. It is also possible that the run-in period did not exclude all prevalent cancers and that some events did not represent first primary cancers. Lastly, we were unable to measure and account for cancer risk factors, such as obesity and smoking, although these have been associated with fatal prostate cancer, not any prostate cancer (61–65); we also could not control for HIV-related factors, such as viral suppression and immune status.

This study was conducted among Medicaid beneficiaries enrolled in 14 US states over the period of 2001-2015. Approximately 40% of PLWH in the US are enrolled in Medicaid (45); thus, the results of this analysis reflect the experience of a significant proportion of men with HIV in the US. Our study benefits from comparing men with HIV to men without HIV who are otherwise similar with respect to socioeconomic characteristics and access to care, complementing prior congruent reports of lower incidence (8–14), some of which were unable to make such direct comparisons. We were also able to estimate differences in prostate cancer incidence by HIV status stratified by race-ethnicity and identified heterogeneity in this association not previously reported. These results extend our understanding of prostate cancer in men with HIV to the Medicaid-insured population; however, existing evidence remains insufficient to identify underlying causes and potential targets of intervention to address differences in prostate cancer incidence and disparities in outcomes by HIV status. Further investigation, with particular emphasis on stage and grade at diagnosis, may provide sufficient evidence to support or exclude hypotheses explaining the differences by HIV status related to prostate cancer screening and detection, as well as biological mechanisms of prostate cancer development.

## Data Availability

The data that support the findings of this study are available from Research Data Assistance Center (ResDAC). Restrictions apply to the availability of these data, which were used under license for this study.

## Author statement

The data that support the findings of this study are available from Research Data Assistance Center (ResDAC). Restrictions apply to the availability of these data, which were used under license for this study. The Johns Hopkins Bloomberg School of Public Health Institutional Review Board determined that this study represented a secondary analysis of existing Medicaid claims data and that it met criteria for exempt status. The authors have no conflicts of interest to disclose.

## Acknowledgements

This work was supported in part by NIH grants R01 CA250851, U01 AI069918, P30 CA006973, and by a 2018 and 2021 development grant from the Johns Hopkins University Center for AIDS Research, an NIH-funded program (1P30AI094189), which is supported by the following NIH Co-Funding and Participating Institutes and Centers: NIAID, NCI, NICHD, NHLBI, NIDA, NIA, NIGMS, NIDDK, NIMHD. F Pirsl was supported by the NCI Cancer Epidemiology, Prevention, and Control Grant (T32 CA0093140). This content does not necessarily represent the official views of the NIH and is the sole responsibility of the authors.

