## Supplemental Materials for "Incidence of prostate cancer in Medicaid beneficiaries with and without HIV in 2001-2015 in 14 states"

Supplement

Supplemental Methods

Variables of interest were identified using ICD-9 (International Classification of Diseases, Ninth Revision) codes. The variables, including specific diagnoses used to define them and codes are listed in the table below:

| Variable | Code type(s) | Code(s) | | Definition | | Date |
| --- | --- | --- | --- | --- | --- | --- |
| HIV | | | | | | |
|  | ICD-9 | 042, 042.X, 079.53, V08 | | 1 inpatient, 2 long-term care, 2 other-nong drug claims within one-year period | | Service date of  first claim |
| Prostate cancer | | | | | | |
|  | ICD-9 | 185.X, V10.46 | | Run-in period (exclusion):  1 claim | | Service date of claim |
|  |  |  |  | Post-run-in period (censoring): 2 claims | | Service date of  first claim |
| Other cancer |  |  | |  | |  |
|  | ICD-9 | 140.XX-172.XX, 174.XX-184.XX, 186.XX-195.8X, 200.XX-208.XX, 238.6 | | Run-in period (exclusion):  1 claim | | Service date of claim |
|  |  |  |  | Post-run-in period (censoring): 1 claim | | Service date of claim |
| Comorbid conditions | | |  | |  | |
| Myocardial infarction | | |  | |  | |
|  | ICD-9 | 410.XX, 412.XX | | 1 claim | | Service date of claim |
| Congestive heart failure | | |  | |  | |
|  | ICD-9 | 398.91, 402.01, 402.11, 402.91, 404.01, 404.03, 404.11, 404.13, 404.91, 404.93, 425.4-425.9, 428.XX | | 1 claim | | Service date of claim |
| Peripheral vascular disease | | |  | |  | |
|  | ICD-9 | 093.0, 437.3, 440.XX-441.XX, 443.1X, 443.9X, 447.1, 557.1, 557.9, V34.4 | | 1 claim | | Service date of claim |
| Cerebrovascular disease | | |  | |  | |
|  | ICD-9 | 362.34, 430.XX-438.XX | | 1 claim | | Service date of claim |
| Dementia | | |  | |  | |
|  | ICD-9 | 290.XX, 294.1X, 331.2 | | 1 claim | | Service date of claim |
| Chronic pulmonary disease | | |  | |  | |
|  | ICD-9 | 416.8, 416.9, 490.XX-505.XX,  506.4, 508.1, 508.8 | | 1 claim | | Service date of claim |
| Rheumatic disease | | |  | |  | |
|  | ICD-9 | 446.5, 710.XX-710.4X, 714.XX-714.2X, 714.81, 714.89, 725.XX | | 1 claim | | Service date of claim |
| Peptic ulcer disease | | |  | |  | |
|  | ICD-9 | 531.XX-534.XX | | 1 claim | | Service date of claim |
| Mild liver disease | | |  | |  | |
|  | ICD-9 | 070.22, 070.23, 070.32, 070.33, 070.44, 070.54, 070.6, 070.9, 570.XX-571.XX, 573.3, 573.4, 573.8, 573.9, V42.7 | | 1 claim | | Service date of claim |
| Moderate to severe liver disease | | |  | |  | |
|  | ICD-9 | 456.0X-456.2X, 572.0X-572.8X | | 1 claim | | Service date of claim |
| Diabetes without chronic complications | | |  | |  | |
|  | ICD-9 | 250.0X-250.3X, 250.8, 250.9 | | 1 claim | | Service date of claim |
| Diabetes with chronic complications | | | | | | |
|  | ICD-9 | 250.4X-250.7X | | 1 claim | | Service date of claim |
| Hemiplegia or paraplegia | | | | | | |
|  | ICD-9 | 334.1, 344.0X-344.6X, 344.9 | | 1 claim | | Service date of claim |
| Renal disease | | | | | | |
|  | ICD-9 | 403.01, 403.11, 403.91, 404.02, 404.03, 404.12, 404.13, 404.92, 404.93, 582.XX-583.7X, 585.XX, 586.XX, 588.0, V42.0, V45.1XX, V56.XX | | 1 claim | | Service date of claim |

Supplemental Results

Medicaid administrative claims data quality is known to vary by state and it is possible for fidelity of some study measures, particularly those not related to billable services, to have differed by state. Most notably, we found reporting of dates of death to be sparse or to appear to be performed in batches sporadically over time in several states and to be minimal across the study period in California. Sensitivity analysis among men aged 50-64 showed estimates obtained from Fine and Gray models were robust to omission of California (shown below). Models omitting California were limited to men aged 50-64 as events predominantly occurred in this age group and these men were at highest risk of death.

|  | All states | | Omitting California |
| --- | --- | --- | --- |
|  | csHR (95% CI) | sdHR (95% CI) | sdHR (95% CI) |
| Overall |  |  |  |
| No HIV | 1.00 (reference) | 1.00 (reference) | 1.00 (reference) |
| HIV | 0.89 (0.80, 0.99) | 0.85 (0.77, 0.95) | 0.84 (0.75, 0.95) |
| NHW, NHB, Hispanic |  |  |  |
| No HIV | 1.00 (reference) | 1.00 (reference) | 1.00 (reference) |
| HIV | 0.85 (0.76, 0.96) | 0.82 (0.73, 0.92) | 0.81 (0.67, 0.97) |
| Non-Hispanic white |  |  |  |
| No HIV | 1.00 (reference) | 1.00 (reference) | 1.00 (reference) |
| HIV | 1.15 (0.88, 1.50) | 1.12 (0.86, 1.46) | 1.22 (0.91, 1.63) |
| Non-Hispanic Black |  |  |  |
| No HIV | 1.00 (reference) | 1.00 (reference) | 1.00 (reference) |
| HIV | 0.81 (0.70, 0.94) | 0.77 (0.67, 0.89) | 0.78 (0.67, 0.90) |
| Hispanic |  |  |  |
| No HIV | 1.00 (reference) | 1.00 (reference) | 1.00 (reference) |
| HIV | 0.80 (0.61, 1.06) | 0.78 (0.59, 1.02) | 0.75 (0.56, 1.00) |

All models adjusted for age (time metric), race-ethnicity, state of residence, enrollment year, and comorbid conditions; model estimates labeled ‘All states’ also presented in Table 3; abbreviations: CI-confidence interval, csHR-cause-specific hazard ratio, NHB-non-Hispanic Black, NHW-non-Hispanic white, sdHR-sub-distribution hazard ratio

Supplemental Tables

Supplemental Table 1: Cause-Specific Hazard Ratios of Prostate Cancer by HIV Status in Men Aged 18-39

|  | Cases | Person-time (PY) | IR^a^ | Beneficiaries | Crude^b^ | Adjusted^c^ |
| --- | --- | --- | --- | --- | --- | --- |
|  |  |  |  |  | csHR (95% CI) | csHR (95% CI) |
| Overall | | | | | | |
| No HIV | 155 | 12,503,999.50 | 1.24 | 10,238,698 | 1.00 (reference) | 1.00 (reference) |
| HIV | 7 | 74,637.90 | 9.38 | 42,864 | 4.41 (2.06, 9.47) | 2.02 (0.93, 4.39) |
| NHW, NHB, Hispanic | | | | | | |
| No HIV | 145 | 10,911,210.25 | 1.33 | 8,905,848 | 1.00 (reference) | 1.00 (reference) |
| HIV | 6 | 69,265.88 | 8.66 | 38,727 | 3.80 (1.67, 8.65) | 1.81 (0.79, 4.18) |
| Non-Hispanic white | | | | | | |
| No HIV | 47 | 4,263,012.50 | 1.10 | 3,272,638 | 1.00 (reference) | 1.00 (reference) |
| HIV | 2 | 17,800.04 | 11.24 | 11,241 | 5.45 (1.31, 22.75) | 4.15 (0.99, 17.43) |
| Non-Hispanic Black | | | | | | |
| No HIV | 62 | 2,502,800.75 | 2.48 | 1,860,913 | 1.00 (reference) | 1.00 (reference) |
| HIV | 4 | 32,951.99 | 12.14 | 17,951 | 2.75 (0.99, 7.64) | 1.81 (0.64, 5.10) |
| Hispanic | | | | | | |
| No HIV | 36 | 4,145,397.00 | 0.87 | 3,772,297 | 1.00 (reference) | 1.00 (reference) |
| HIV | 0 | 18,513.85 | 0.00 | 9,535 | NE | NE |

^a^per 100,000 person-years, ^b^adjusted for age (time metric), ^c^adjusted for age (time metric), race-ethnicity, state of residence, enrollment year, and comorbid conditions; abbreviations: CI-confidence interval, csHR-cause-specific hazard ratio, IR-incidence rate, NE-not estimated, NHB-non-Hispanic Black, NHW-non-Hispanic white, PY-person-years, sdHR-sub-distribution hazard ratio

Supplemental Figures

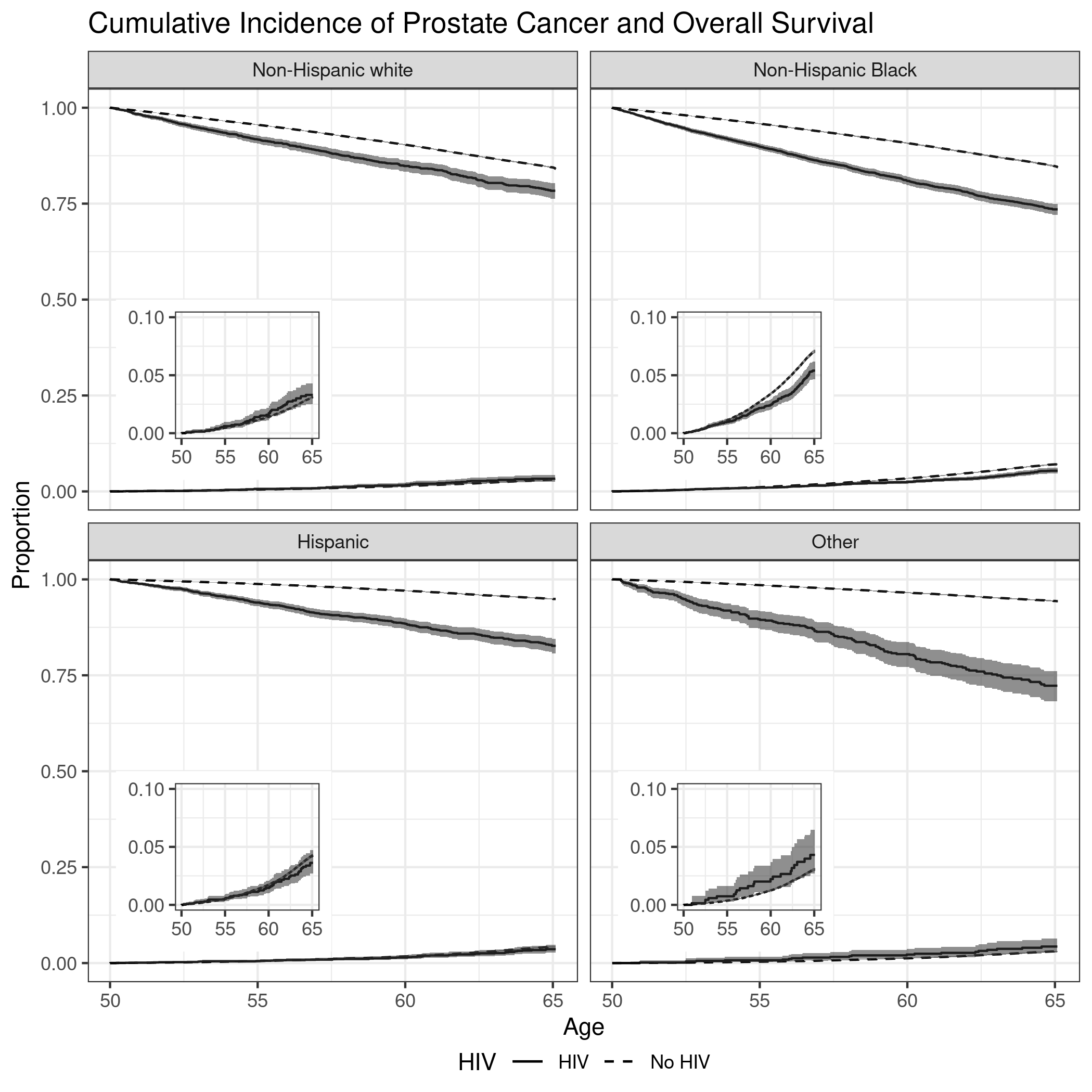

Supplemental Figure 1: Cumulative Incidence of Prostate Cancer and Overall Survival by HIV Status in Men Aged 50-64 Stratified by Race-Ethnicity; insets are magnified curves for cumulative incidence of prostate cancer

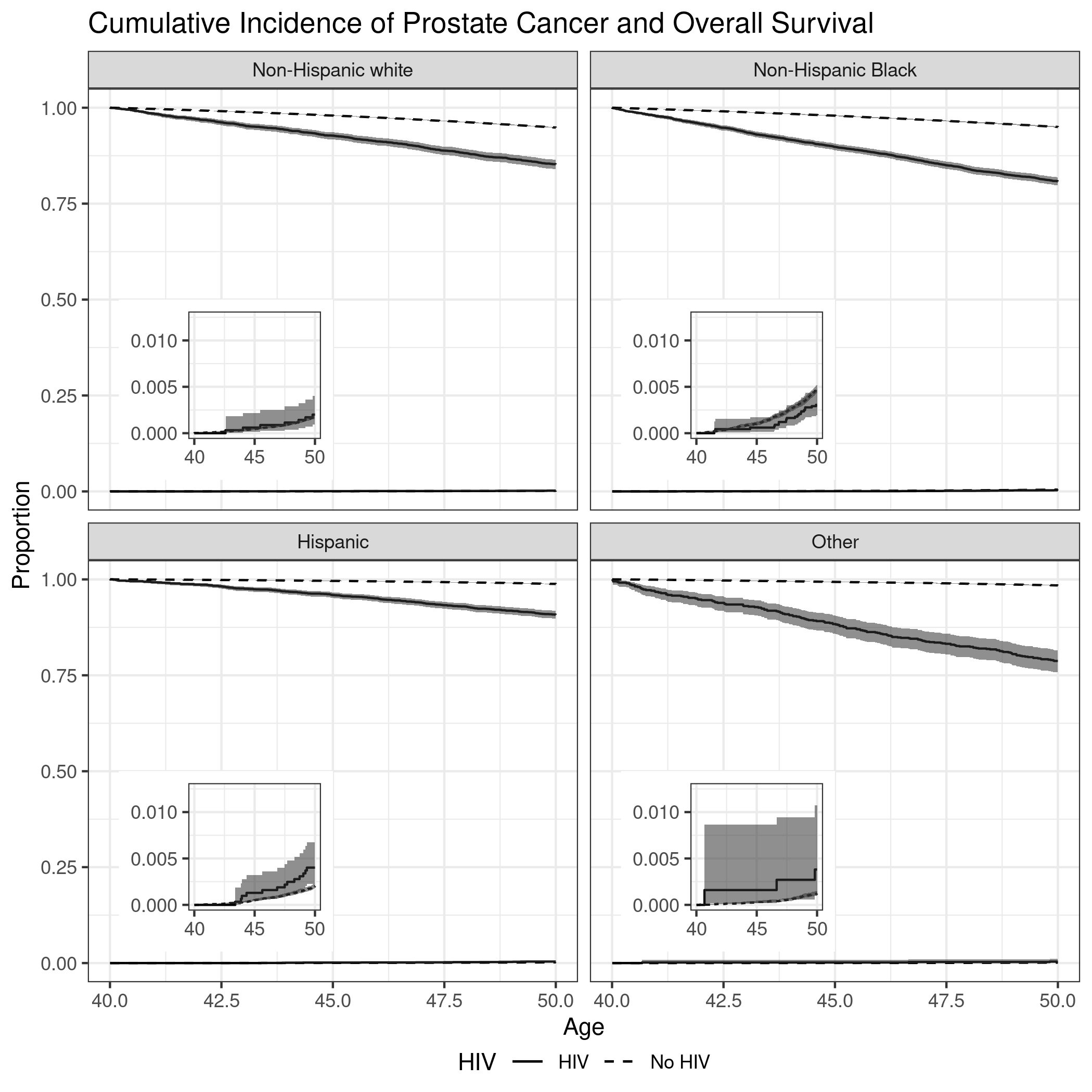

Supplemental Figure 2: Cumulative Incidence of Prostate Cancer and Overall Survival by HIV Status in Men Aged 40-49 Stratified by Race-Ethnicity; insets are magnified curves for cumulative incidence of prostate cancer

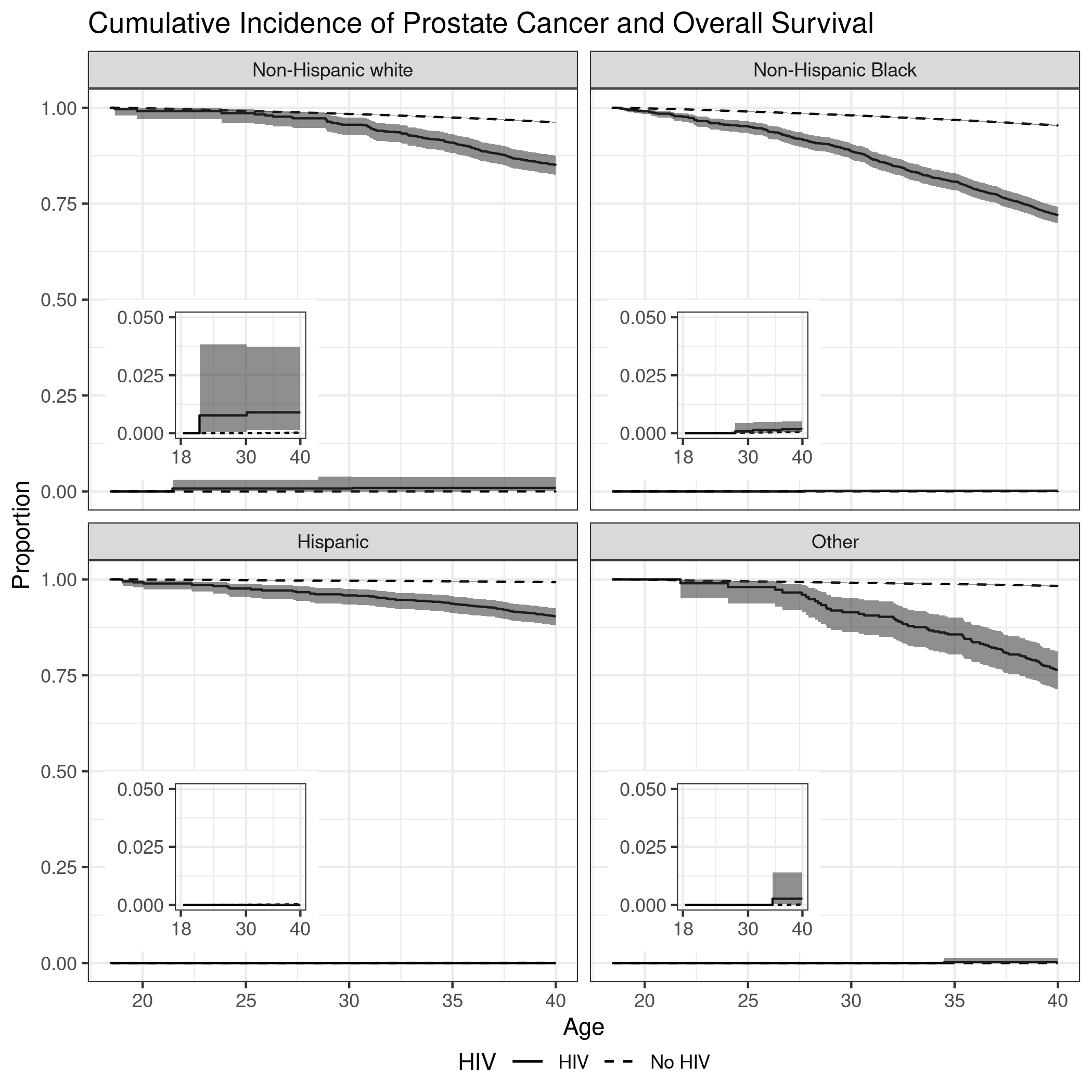

Supplemental Figure 3: Cumulative Incidence of Prostate Cancer and Overall Survival by HIV Status in Men Aged 18-39 Stratified by Race-Ethnicity; insets are magnified curves for cumulative incidence of prostate cancer
